# Misdiagnosis and underdiagnosis of glioma: Systematic review

**DOI:** 10.1101/2025.04.03.25325091

**Authors:** Dariya Ilchenko, Wolfgang Emanuel Zürrer, Amelia Cannon, Marco Piccirelli, Zsolt Kulcar, Sebastian Winklhofer, Benjamin Victor Ineichen

## Abstract

**Background:** Diagnostic errors in gliomas, a major group of brain tumors originating from glial cells, can have severe consequences for patients and healthcare systems. Despite the serious implications of a glioma diagnosis, there is a lack systematic evidence on the frequency of diagnostic errors. This study aimed to assess the prevalence of misdiagnosis and underdiagnosis of gliomas and their potential impact.

**Methods:** We conducted a systematic review of English-language original studies, including those with more than 10 participants, reporting diagnostic errors in gliomas. A search of Medline and Embase was performed from inception until October 28, 2024. We evaluated the proportions of diagnostic errors and their potential consequences, and assessed the risk of bias using a modified Newcastle-Ottawa Scale.

**Results:** Of 1,860 studies screened, 22 met the inclusion criteria. Our analysis indicates that gliomas are frequently underdiagnosed (i.e., the correct diagnosis is missed) and, though less extensively reported, also misdiagnosed (i.e., assigned incorrect diagnoses). Overall, diagnostic errors ranged from 1.9 to 11.6% with a median of 5% (mis-/underdiagnosis: 5%, range 1.9-20%, misgrading: 11.6% range: 0.6-47%). While diagnosing gliomas is complex, evidence suggests that clinical tools such as MRI and pathological methods, including neuroendoscopic biopsy and specific cytology techniques, can achieve high diagnostic accuracy. Diagnostic errors were linked to relevant patient consequences, with both overtreatment and undertreatment commonly reported.

**Conclusion:** Diagnostic errors in gliomas are common and can have serious implications for patients. More rigorous data are needed to better understand the causes of these errors, which is key for reducing misdiagnosis and underdiagnosis in the future.

## Introduction

Gliomas constitute a major group in the 5th edition of the World Health Organization (WHO) classification, forming a diverse group of central nervous system (CNS) tumors that originate from glial cells [1, 2]. These tumors are traditionally classified based on their grade of malignancy and histological type [3]. While most gliomas are known for their diffuse infiltration into surrounding tissues, some exhibit a more defined growth pattern. A notable update in the latest WHO classification is the segregation of diffuse gliomas into adult and pediatric types [4], acknowledging their distinct biological and clinical characteristics. This delineation results in four general groups: adult-type diffuse gliomas, pediatric-type diffuse low-grade gliomas, pediatric-type diffuse high-grade gliomas, and circumscribed astrocytic gliomas [5]. This evolution in classification underscores the complexity and heterogeneity of gliomas, warranting more tailored diagnostic and therapeutic approaches.

The diagnosis of a glioma has a profound impact, not only due to the high degree of malignancy associated with these tumors but also because of their substantial mortality rates [6]. In the United States, between 1975 and 2018, over 60,000 patients received a glioma diagnosis, with the overall mortality rate exceeding 50%. Gliomas account for approximately 30% of all brain and CNS tumors and 80% of all malignant brain tumors [7]. Consequently, an accurate diagnosis of glioma, which includes determining the precise stage of cancer, is key. It influences the treatment options available and, ultimately, the patient’s prognosis [8]. However, achieving an accurate diagnosis is a meticulous task and presents challenges, including the potential for misdiagnosis, misgrading, or underdiagnosis due to a wide array of neuroimaging mimics. And despite the critical impact of such diagnostic errors, there is a notable lack of comprehensive evidence summarizing diagnostic errors for glioma.

Based on this shortcoming, the aim of our study was to conduct a systematic review of glioma diagnostic errors, including misdiagnosis (i.e., being assigned with an incorrect glioma diagnosis), misgrading, and underdiagnosis (i.e., the correct glioma diagnosis is missed) of gliomas. Thus, we included studies reporting errors from complete clinical workups as well as those assessing specific diagnostic methods, such as MRI. With this, our study aims to enhance diagnostic accuracy for gliomas.

## Methods

### Protocol registration

We registered the study protocol in the International prospective register of systematic reviews (PROSPERO, CRD42022333679, https://www.crd.york.ac.uk/PROSPERO/) and used the Preferred Reporting Items for Systematic Reviews and Meta-Analysis (PRISMA) Guidelines for reporting [9].

### Search strategy

We searched for studies from inception to October 28, 2024, in Medline via PubMed and EMBASE via the Ovid interface. See **Table S1** for the search string in each of these databases. In addition, we searched reference lists of eligible publications for additional potentially relevant references.

### Inclusion and exclusion criteria

#### Inclusion criteria

Any studies presenting original findings on glioma misdiagnosis or underdiagnosis comprising ≥ 10 human individuals. We included all clinical study designs, e.g., case series, case-control, and cohort studies.

#### Exclusion criteria

Studies not addressing glioma diagnostic errors, non-English studies, conference abstracts, and publications that reiterated previously reported quantitative data. We excluded (systematic) reviews but retained them as potential sources of additional records.

### Study selection and data extraction

Two reviewers (DI and MP) screened titles and abstracts of studies for their relevance in the web-based application Rayyan [10], followed by full-text screening. Subsequently, we extracted the following data: title, authors, publication year, number of subjects per group, mean/median age and sex of participants, study country, misdiagnosis/underdiagnosis proportion, incorrect and correct diagnoses, potential unnecessary exposure to treatment, duration until correct diagnosis, reasons for diagnostic errors, and potentially associated morbidity. We contacted corresponding authors of included studies via email in case of missing data.

### Risk of bias assessment

Two reviewers (DI and BVI) assessed risk of bias of eligible studies using an adjusted version of the Newcastle-Ottawa scale, considering bias in the selection, comparability, and exposure domain [11].

### Data synthesis and analysis

We summarized findings in narrative fashion and present descriptive statistics for demographic parameters (age and sex), misdiagnosis/underdiagnosis proportions (including categorizations by sex), associated morbidity, and proportions of diagnostic errors in relation to sex.

### Publication bias

We did not assess publication bias, as pre-defined in our study protocol.

## Results

### 1. Eligible publications

In total, 1940 studies were retrieved from our database search. After abstract and title screening, 332 studies were eligible for full-text search. After screening the full text of these studies, 22 studies were included for qualitative synthesis (**Figure 1**).

**Figure 1:**
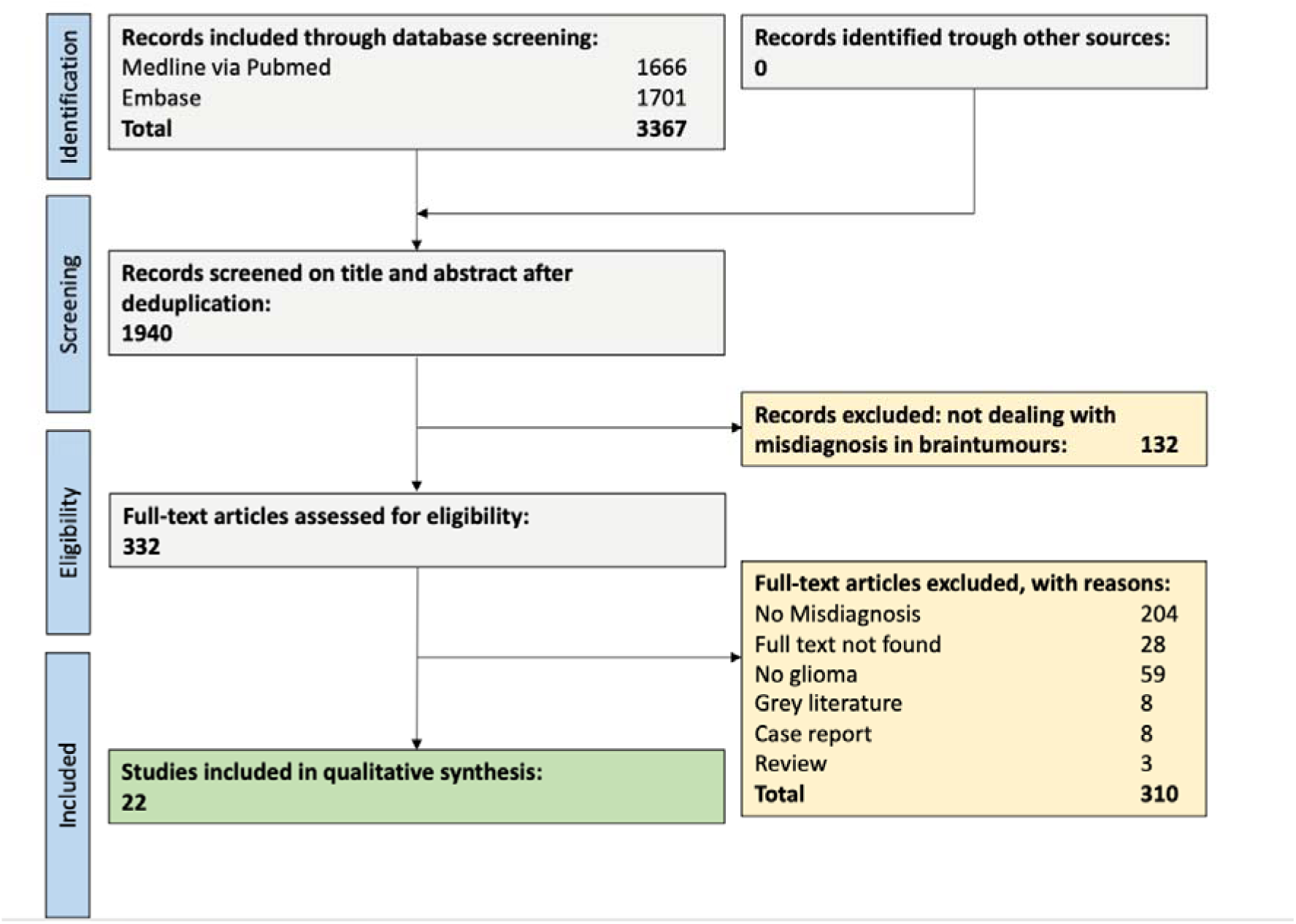
Study flow chart.

### 2. General study characteristics

#### Included publications

Out of all 22 studies, 20 assessed individuals who were underdiagnosed with glioma (**Table 1**). Eleven of these 20 studies also reported cases of glioma misdiagnosis. Most studies (20 studies, 95%) were published after 2000. Studies originated from 12 different countries, with most studies from USA (8 studies) as well as China and Japan (2 studies each).

**Table 1:** See supplementary file.

Overall, the studies included 3624 individuals, with 1871 men (52%) and 1436 women (39%) (sex not reported for the remaining individuals). Most studies included both children and adults (>18 years old; 9 studies). One study included children only [12].

#### Risk of bias assessment

The studies exhibited a low risk of bias in the comparability and exposure domain, represented by their inclusion of cases with definite diagnoses, reporting of alternative diagnoses and case demographics, and clear provision of patient in- and exclusion criteria (**Table S2**). However, there was a high risk of bias in parts of the selection domain (S1) due to their focus only on cases with a definite or suspected glioma diagnosis, rather than assessing diagnostic errors at a population level.

### 3. Overall diagnostic errors

Overall, diagnostic errors ranged from 1.9% in a 2001 study evaluating unclear intraoperative CNS lesions using cytology [13] to 47% in a 2000 study using single-voxel MR spectroscopy [14], with a median of 11.6%. For under-/misdiagnosis, it ranged from 1.9% in a 2001 study evaluating unclear intraoperative CNS lesions using cytology [13] to 20% in a 2000 study on desmoplastic neuroepithelial tumors in childhood [12], with a median of 5%. Misgrading ranged from 0.6% in a 2018 study comparing different intraoperative cytology techniques [15] to 47% in a 2000 study using single-voxel MR spectroscopy [14], with a median of 9%. Focusing on studies from the past five years (4 studies [16–19]), diagnostic errors ranged from 2.8% to 9.1%, with a median of 5.6%. For misgrading, one study reported a rate of 3.9% [16].

### 4. Underdiagnosis of gliomas

Twenty studies assessed individuals who were underdiagnosed with glioma (i.e., missing a correct glioma diagnosis). The most frequently reported diagnostic error was the misgrading of gliomas, e.g., due to incorrect biopsy locations or ambiguous MRI scans, the incorrect assessment of their tissue of origin. The entities diagnosed instead of glioma are reported in **Table 1** and summarized in **Table 2**.

**Table 2:** Misdiagnoses (i.e., being assigned with an incorrect glioma diagnosis) and underdiagnoses for gliomas (i.e., the correct glioma diagnosis is missed).

| Misdiagnosis: Disease incorrectly diagnosed as glioma | Underdiagnosis: glioma is incorrectly diagnosed as |
| --- | --- |
| Ependymoma (anaplastic)<br>Gliosis<br>Lymphoma<br>Medulloblastoma<br>Meningioma<br>Metastasis<br>Necrotic tissue<br>Normal brain tissue<br>Pineocytoma<br>Small round blue cell tumor<br>Stroke | Anaplastic ependymoma<br>Astrogliosis<br>Benign teratoma<br>Cavernoma<br>Congenital glial cyst<br>Demyelinating disease (multiple sclerosis)<br>Dysembryoplastic neuroepithelial tumor<br>Ependymoma<br>Gliosarcoma<br>Hemangiopericytoma<br>Lymphoma<br>Meningioma<br>Metastatic (lung) carcinoma<br>Metastatic carcinoma<br>PNET (Primitive neuroectodermal tumor)<br>Stroke/hemorrhagic infarction |

#### Neuroimaging

The early work by Kendall already in 1979 in the UK highlighted the limitations of relying solely on CT scans for glioma diagnosis, reporting common misdiagnosis and misgrading of tumours [20]. They reported that 1.5% of gliomas were missed on the initial scan and 6.5% of the gliomas found were misgraded. Two more recent studies from 2018 and 2019 explored MRI features helpful at distinguishing gliomas from other brain pathologies [21], such as autoimmune limbic encephalitis [17]. These MRI features included among others mass effects, edema, and contrast enhancement patterns. Another 2021 study compared stroke with gliomas emphasizing that gliomas can also mimic strokes [18]. Moreover, the accuracy of magnetic resonance spectroscopy (MRS) in diagnosing gliomas was examined, and highlighting MRS’s practicality when guided by multimodal MR imaging [22], yet proper placement of the volume of interest is important for the diagnostic accuracy of this approach [14]. Concretely, if positioned at the edge of the tumour, the spectra obtained are more accurate compared with placement in the lesion center. It has also been suggested that measures of discordant relative cerebral blood volume, in conjunction with histopathology, may help to identify patients at higher risk for glioma undergrading [23].

#### Biopsy and histopathology

An early Polish study assessed the advantages and limitations of the intraoperative smear technique for diagnosing brain tumours including gliomas [24]. Findings indicated the technique’s rapidity, simplicity, and relatively high diagnostic efficacy as its primary benefits. Notably, even very small and soft specimens proved suitable for this method, an advantage for surgeries involving tumours in functionally significant areas of the brain. This finding received further validation from two subsequent studies [13, 25]. However, the need for re-evaluation and comparison with final histological examinations was also emphasized [26], pointing out its importance for accurately grading gliomas. Building on this, a comprehensive Japanese study investigated the diagnostic effectiveness of combining squash and touch preparation cytology [15]. This combined approach showcased near-perfect sensitivity and specificity, achieving 95% overall diagnostic accuracy. It was particularly effective in diagnosing brain tumours susceptible to smearing artifacts and certain metastatic malignancies. Another study confirmed these findings, uncovering a diagnostic accuracy of almost 90% to diagnose gliomas [16].

A 2003 study compared the accuracy of MR-guided stereotactic brain biopsies in gliomas with the findings from subsequent specimen resections, showing a high agreement rate of 96% [27]. Building on this work, another study investigated the diagnostic challenges and risks associated with stereotactic biopsy when evaluating intra-axial brain mass lesions [28]. Both studies found that inaccuracies in biopsy procedures could complicate the process of establishing a correct diagnosis. Another study compared the diagnostic reliability of stereotactic biopsy against diagnoses made using clinical data and neuroimaging techniques in 200 patients with brain tumours: it showed that stereotactic biopsies resulted in incorrect diagnoses in 7% of cases, suggesting that in instances where a clear presumptive diagnosis exists, clinical assessments and neuroimaging findings may alone be sufficient [29]. Also, MRI findings have been compared with findings from stereotactic brain biopsy results, particularly in distinguishing between high and low-grade gliomas [30]. The study concluded that stereotactic biopsy plays an important role in grading when MRI findings suggest low-grade gliomas. Finally, an international multicenter study analyzing the accuracy and safety of neuroendoscopic biopsies in brain tumors compared such neuroendoscopic biopsies with results from open surgery [31]. The study reported that neuroendoscopic biopsies achieve an accurate diagnosis in 90% of cases. In around 18% of the cases, there was disagreement on the pathology between the neuroendoscopic biopsy and the pathology from the open surgery. In total, 12% of the total cases would have been mismanaged based on the findings of the neuroendoscopic biopsy.

#### Population-based approaches

A large study from 2000 assessed diagnostic discrepancies in gliomas among more than 450 patients in the San Francisco Bay area [32]. The findings showed that glioma misdiagnosis rates were substantially higher in patients from community hospitals compared to those treated in academic centers. Discordant diagnoses occurred more commonly in community hospitals versus academic centers (26% versus 12%). Among the 105 discordant cases, 17 (16%) were deemed clinically meaningful, suggesting the discrepancy could have a relevant impact on patient management and/or prognosis. Following a similar line of investigation, a study of rare childhood brain tumors found that preoperative biopsies led to misdiagnoses in 2 out of 10 cases, thereby questioning the necessity of such biopsies in certain situations [12].

### 5. Misdiagnosis of gliomas

Two studies reported cases of glioma misdiagnosis (i.e., incorrectly assigning a glioma diagnosis), partly overlapping with cases of underdiagnosis. The most frequently reported misdiagnoses were non-glioma tumours of the CNS as well as stroke and demyelinating disorders (**Tables 1 and 2**).

#### Misdiagnosis by modality

In 2009 a German study found that dysembrioplastic neuroepithelial tumors (DNTs) are occasionally misdiagnosed as gliomas [33]. They investigated whether the presence of glial nodules lead to a misdiagnosis of glioma. They found that the glioneural element can be identified on MRI, hence it should be used to support the diagnosis of DNT to minimize misdiagnosis of DNT as glioma.

The study of Yu et al from 2023 aimed to assess the diagnostic efficacy of non-enhancing tumor volume, apparent diffusion coefficient and arterial spin labeling in distinguishing atypical primary central nervous system lymphoma (PCNSL) from glioblastoma (GBM) [19]. The findings emphasize the importance of multiparametric assessment in clinical decision-making to prevent unnecessary surgical interventions and ensure accurate diagnosis.

### 6. Consequences of misdiagnosis

Among the most impactful consequences reported was inappropriate treatment. For example, some patients received unnecessary chemotherapy or radiation therapy [12, 26], while others experienced delayed treatment due to late diagnosis, postponing proper intervention [18, 21]. Interestingly, Aldape’s study highlighted that although there were more upgrades than downgrades in glioma grading, the downgrades were clinically more relevant [32]. Inaccurate diagnosis of atypical primary central nervous system lymphomas (PCNSL) could lead to unnecessary surgical resection, exposing patients to avoidable surgical risks [19]. Vaquero also noted that diagnostic errors significantly impacted prognosis and therapeutic management, with some patients succumbing to tumor progression after being incorrectly treated for high-grade astrocytoma [29].

Another critical consequence of delayed diagnosis was the postponement of appropriate treatment. In Sasagawa’s study, a patient misdiagnosed with stroke waited over two months before receiving the correct diagnosis of glioblastoma, enduring an unnecessary stroke workup [18]. Similarly, Mallucci’s study documented a case where an erroneous diagnosis of malignant grade 4 astrocytoma resulted in perioperative death, as the patient underwent preoperative chemotherapy without effect [12].

## Discussion

### Main findings

The aim of our study was to systematically review the glioma diagnostic errors, including misdiagnosis, misgrading, and underdiagnosis. Our analysis shows that gliomas are frequently underdiagnosed (i.e., the correct diagnosis is missed) and although less extensively documented, they are also misdiagnosed (i.e., being assigned with incorrect diagnoses). Despite the complexity of diagnosing gliomas, the evidence suggests that clinical tools such as MRI and pathological methods, including neuroendoscopic biopsy and specific cytology techniques, can be employed to achieve high sensitivity and specificity for glioma diagnosis. Importantly, diagnostic errors are associated with relevant consequences for the patients, with both overtreatment and undertreatment frequently reported in the literature.

### Findings in the context of existing evidence

Gliomas are frequently underdiagnosed, meaning the correct diagnosis is missed, along with frequent misgrading, supported by a substantial evidence base. On the other hand, we also find that gliomas are often misdiagnosed, though this is based on a smaller body of evidence. Notably, a large population-based study reported higher rates of diagnostic errors in community hospitals compared to academic centers [32], likely reflecting the greater expertise and more advanced diagnostic tools available at academic institutions [34]. Similar patterns have been observed in other neurological conditions, such as stroke [35].

Common differential diagnoses to consider include other primary brain tumors like ependymoma, lymphoma, medulloblastoma, glioblastoma, meningioma, oligodendroglioma, astrocytoma, PNET, and DNT, as well as metastases. Stroke and gliosis can also be mistaken for gliomas. Beyond surgical methods such as neuroendoscopic biopsies and cytology techniques, MRI plays a crucial role in distinguishing gliomas from their common mimics, like brain metastases [36]. However, this often requires more advanced techniques such as radiomics, an emerging method that analyzes the intensity, shape, and texture of medical images—features invisible to the naked eye—to identify tumors [37]. It is important to note, however, that the quality of studies testing potential methods to enhance diagnostic accuracy is relatively low, as highlighted by a recent Cochrane systematic review [38].

Although the gold standard for glioma diagnosis remains histological work-up [39]. Nevertheless, accurate glioma diagnosis also requires integration of histological, immunohistochemical, and molecular findings with imaging and demographic data. Key molecular markers such as IDH mutational status and 1p/19q codeletion are essential for classification and prognostication [39]. For instance, molecular alterations like those found in glioblastoma and diffuse midline gliomas can predict poor outcomes despite lower-grade histology, underscoring the importance of molecular profiling in optimizing treatment strategies [40].

Modern diagnostic approaches also leverage advancements in metabolic imaging. These offer non-invasive approaches to improve diagnostic accuracy and patient management [41]. Techniques such as PET and MR spectroscopy leverage cancer-specific metabolic phenomena to potentially detect unique tumor signatures [42]. Also, artificial intelligence (AI) is emerging as an increasingly important tool in neuro-oncology, contributing to lesion detection, segmentation, and molecular marker identification [43]. Machine learning and deep learning algorithms have shown potential in improving diagnostic precision and workflow efficiency. For example, combining stimulated Raman histology with deep learning has enabled accurate intraoperative differentiation of CNS lesions within minutes [44].

The consequences of diagnostic errors can be severe, involving both overtreatment and undertreatment. Overtreatment, such as unnecessarily exposing a patient to chemotherapy and/or radiotherapy, can lead to serious short- and long-term effects, including neutropenia, opportunistic infections, and, in the worst cases, secondary cancers over time [45–47]. On the other hand, undertreatment can be equally dangerous, as prompt treatment following a glioma diagnosis is a key predictor of better outcomes [48, 49]. Overall, medical errors rank as the third leading cause of death in the USA [50]. Therefore, accurate glioma diagnosis demands a diligent workup to minimize these risks [49].

### Limitations

Our study has several notable limitations. First, assessing the extent of diagnostic error for any disease is methodologically challenging and, by nature, retrospective [51]. As a result, the actual rates of misdiagnosis or underdiagnosis of gliomas might be overestimated in our systematic review, partly due to the lack of reported misdiagnosis rates in the general population. Second, there was high heterogeneity on reporting and methodology in the included publications. Third and most importantly, the evolution and revision of glioma diagnostic criteria over time may affect the consistency and comparability of misdiagnosis rates across studies. As these criteria change, they may influence both the prevalence and nature of misdiagnoses, complicating direct comparisons of data from different time periods in our systematic review. Along these lines, advancements in medical diagnostics and the adoption of modern techniques likely improve diagnostic accuracy over time. While we attempted to account for this through a sensitivity analysis of studies from the last five years, this temporal factor still adds complexity to the interpretation of our findings.

### Conclusions

In conclusion, our study provides systematic evidence of the high rates of misdiagnosis and underdiagnosis of gliomas, with potentially serious consequences for affected individuals. More rigorous data is essential to better understand the underlying causes of these diagnostic errors, which will be critical for reducing diagnostic errors in the future. A diligent clinical work including modern neuroimaging methods is warranted to enhance diagnostic accuracy for gliomas.

## Declarations

### Ethical approval and consent to participate

Not applicable.

### Consent for publication

Not applicable.

### Availability of data and materials

The dataset supporting the conclusions of this article is included within the article and its additional files.

### Author contributions

Conception and design of study: DI, WEZ, AEC, MP, BVI; acquisition of data: DI, BVI; analysis of data: DI; drafting the initial manuscript: DI, BVI; all authors critically revised the paper draft.

## Acknowledgments

We thank Sergej Prokofiev for help with data analysis.

