## Supplementary data for "Misdiagnosis and underdiagnosis of glioma: Systematic review"

**Table S1**: Search string

(brain neoplasms [MeSH])

AND

(misdiagnosis OR "diagnostic error" OR misdiagnosed OR misattributed OR “wrongly diagnosed” OR “incorrectly diagnosed” OR “wrong diagnosis” OR "diagnostic delay" OR "delayed diagnosis")

**Table S2:** Risk of bias assessment of the included studies.

|  | **Title** | **S1** | **S2** | **S3** | **C1** | **C2** | **E1** |
| --- | --- | --- | --- | --- | --- | --- | --- |
| Aldape, 2000 | Discrepancies in diagnoses of neuroepithelial neoplasms: the San Francisco Bay Area Adult Glioma Study | 1 | 1 | 1 | 1 | 1 | 1 |
| Callovini, 2008 | Is it appropriate to redefine the indication for stereotactic brain biopsy in the MRI Era? Correlation with final histological diagnosis in supratentorial gliomas | 0 | 1 | 1 | 1 | 1 | 1 |
| Campos, 2009 | Simple and complex dysembryoplastic neuroepithelial tumors (DNT) variants: clinical profile, MRI, and histopathology | 0 | 1 | 1 | 1 | 1 | 1 |
| Constantini, 2013 | Safety and diagnostic accuracy of neuroendoscopic biopsies: an international multicenter study | 0 | 1 | 1 | 1 | 1 | 1 |
| Ghosal, 2011 | Smear preparation of intracranial lesions: a retrospective study of 306 cases | 1 | 1 | 1 | 1 | 1 | 1 |
| Hamasaki, 2018 | Intraoperative Squash and Touch Preparation Cytology of Brain Lesions Stained with H+E and Diff-Quik™: A 20-Year Retrospective Analysis and Comparative Literature Review | 0 | 1 | 1 | 1 | 1 | 1 |
| Heper, 2005 | An analysis of stereotactic biopsy of brain tumors and nonneoplastic lesions: a prospective clinicopathologic study | 0 | 1 | 1 | 1 | 1 | 1 |
| Kendall, 1979 | Difficulties in diagnosis of supratentorial gliomas by CAT scan | 0 | 1 | 1 | 0 | 0 | 1 |
| Kraus, 2000 | Long-term survival of glioblastoma multiforme: importance of histopathological reevaluation | 0 | 1 | 1 | 1 | 1 | 1 |
| Maldonado, 2018 | Features of diffuse gliomas that are misdiagnosed on initial neuroimaging: a case control study | 0 | 1 | 1 | 1 | 1 | 1 |
| Mallucci, 2000 | The management of desmoplastic neuroepithelial tumours in childhood | 0 | 1 | 1 | 1 | 1 | 1 |
| McCullough, 2018 | Preoperative relative cerebral blood volume analysis in gliomas predicts survival and mitigates risk of biopsy sampling error | 0 | 1 | 1 | 1 | 1 | 1 |
| McGirt, 2003 | MRI-guided stereotactic biopsy in the diagnosis of glioma: comparison of biopsy and surgical resection specimen | 0 | 1 | 1 | 0 | 1 | 1 |
| Obeidat, 2019 | Accuracy of Frozen-Section Diagnosis of Brain Tumors: An 11-Year Experience from a Tertiary Care Center | 0 | 1 | 1 | 1 | 1 | 1 |
| Ricci, 2000 | Effect of voxel position on single-voxel MR spectroscopy findings | 0 | 1 | 1 | 1 | 1 | 1 |
| Sasagawa, 2021 | Stroke Mimics and Chameleons from the Radiological Viewpoint of Glioma Diagnosis | 0 | 1 | 1 | 1 | 1 | 1 |
| Savargaonkar, 2001 | Utility of intra-operative consultations for the diagnosis of central nervous system lesions | 0 | 1 | 1 | 0 | 1 | 1 |
| Slowinski, 1999 | Smear technique in the intra-operative brain tumor diagnosis: its advantages and limitations | 0 | 1 | 1 | 1 | 1 | 1 |
| Vaquero, 2000 | Stereotactic biopsy for brain tumors: is it always necessary? | 0 | 1 | 1 | 1 | 1 | 1 |
| Yao, 2016 | The clinical utility of multimodal MR image-guided needle biopsy in cerebral gliomas | 0 | 1 | 1 | 1 | 1 | 1 |
| Zoccarato, 2019 | Conventional brain MRI features distinguishing limbic encephalitis from mesial temporal glioma | 1 | 1 | 1 | 1 | 1 | 1 |
| Yu, 2022 | Atypical primary central nervous system lymphoma and glioblastoma: multiparametric differentiation based on non-enhancing volume, apparent diffusion coefficient, and arterial spin labeling | 0 | 1 | 1 | 1 | 1 | 1 |

With 10 or more included human subjects according to the Newcastle-Ottawa scale for nonrandomized studies. S1-S3: Selection domain, C1: comparability domain, E1: exposure domain. Studies are listed in alphabetical order. 0 = no, 1 = yes.
