## Supplementary material for "Misdiagnosis and underdiagnosis of glioma: Systematic review": Table 1

**Table 1**: Studies reporting on diagnostic errors in glioma.

|  | **Country and patient selection** | **Demographics** | **Misdiagnosis rate** | **Initial (wrong) diagnosis/grade** | **Final (correct) diagnosis/grade if underdiagnosed** | **Final (correct) diagnosis/grade if misdiagnosed** |
| --- | --- | --- | --- | --- | --- | --- |
| Kendall, 1979 | UK  “The clinical and pathological records of all patients in the National Hospital for Nervous Diseases with a diagnosis of supratentorial glioma during the period of 1976 and 1977 were reviewed.” | N = 314 | **Diagnostic errors 40/314 (12.7%)**  **Total: 40/314 (12.7%)** | - Meningioma  - Metastasis  - Inflammation  - Cerebral infarction  - Intracranial haemorrhage  - Vascular malformation  - Unspecified mass  - Normal scan |  | Glioma |
| Slowinski, 1999* | Poland  “Specimens obtained via open biopsy during craniotomy performed in 217 patients with a pre-operative diagnosis of brain tumor” | N = 217  Age range 16-76  M:F 110:107 | **Diagnostic errors 6/217 (2.8%)**  **Misgraded 8/217 (3.7%)**  **Total: 14/217 (6.5%)** | - Glioma benignum  - Normal brain tissue  - Glioma malignum  - Astrocytoma  - Glioblastoma multiforme | - Glioma malignum  - Astrocytoma  - Glioma benignum | - Gliosis  - Carcinoma metastaticum |
| Aldape, 2000* | USA  “All histologically confirmed incident cases of glioma and medulloblastoma in adults age 20 and older diagnosed between August 1991 and April 1994” | N = 457  Age mean 54.4  M:F 260:197 | **Diagnostic errors 10/457 (2.1%)**  **Misgraded 95/457 (20.8%)**  **Total: 105/457 (22.9%)** | - Glioblastoma  - Astrocytoma/Gliosarcoma  - Glioblastoma/ Astroblastoma  - Glioblastoma  - Anaplastic astrocytoma  - Malignant glioma |  | - Astrocytoma  - Glioblastoma  - Oligodendro glioma  - Oligoastro cytoma  - Juvenile pylocystic astrocytoma  - Anaplastic astrocytoma |
| Kraus, 2000* | Germany  “Histological preparations of 106 patients with malignant gliomas were retrieved from the archives of four participating institutions” | N = 106  Age mean 52  M:F 63:43 | **Misgraded 14/106 (13.2%)**  **Total: 14/106 (13.2%)** | Glioblastoma multiforme grade IV | - Anaplastic oligodendroglioma Grade III  - Anaplastic oligoastrocytoma Grade III  - Anaplastic astrocytoma Grade III  - Anaplastic pilocytic astrocytoma Grade III |  |
| Mallucci, 2000* | France  “Between 1985 and 1996, ten children were treated for paediatric brain tumours in our institution; the diagnoses were DACI, DIGG and PXA” | N = 10  Age median 9.5 months  M:F 7:3 | **Diagnostic errors 2/10 (20%)**  **Total: 2/10 (20%)** | Malignant grade 4 Astrocytoma |  | - Desmoplastic astrocytoma of infancy (DACI)  - Desmoplastic infantile ganglioglioma (DIGG)  - Pleomorphic xanthoastrocytoma (PXA) |
| Ricci, 2000 | USA  “We retrospectively reviewed 50 consecutive MR imaging and single-voxel proton MR spectroscopy studies in 43 patients with intraaxial brain tumors referred for imaging over an 8-month period by the neuro-oncology and neurosurgery services” | N = 17  Age mean 55  M:F 9:8 | **Misgraded 8/17 (47%)**  **Total: 8/17 (47%)** | - Primary and recurrent glioblastomas - primary anaplastic astrocytoma  - Metastasis | - Primary/recurrent glioma |  |
| Vaquero, 2000 | Spain  “Between 1991 and 1996, 200 patients were subjected to stereotactic biopsy of a presumed brain tumor at the university hospital in Madrid, Spain.” | N = 200  Age mean 48  M:F 134:66 | **Diagnostic errors 19/180 (10.5%)**  **Total: 19/180 (10.5%)** | - Metastasis  - Primary cerebral lymphoma  - High grade astrocytoma  - Low grade astrocytoma |  | - High grade astrocytoma  - Low grade astrocytoma  - Lung carcinoma metastasis  - Pleomorphic xanthoastrocytoma |
| Savargaonkar, 2001* | USA  “From January 1997-June 1999, 103 consecutive Iop consultations for CNS lesions were performed.” | N = 103  Age median NA  M:F 60:43 | **Diagnostic errors 2/103 (1.9%)**  **Total: 2/103 (1.9%)** | - Metastatic tumor  - Small round blue cell tumor |  | - Glioblastoma multiforme  - Anaplastic small cell astrocytoma |
| McGirt, 2003* | USA  “43 patients were identified that had undergone MRI-guided stereotactic brain biopsy followed by open craniotomy at Duke University Medical Center between July 1997 and June 2000.” | N = 43  Age NA  M:F 24:19 | **Misgraded 13/43 (30.2%)**  **Total: 13/43 (30.2%)** | - Anaplastic astrocytoma  - Necrosis/gliosis | Upgraded:  - Astrocytoma III - GBM  - necrosis/gliosis - Astrocytoma III  - Necrosis/gliosis – Glioblastoma multiforme |  |
| Heper, 2005* | Turkey  “Between 1995 and 2003, 130 cases with intra-axial brain lesions underwent SB (stereotactic biopsy) at the Department of Neurosurgery, School of Medicine, Ankara University.” | N = 130  Age mean 46  M:F 85:45 | **Diagnostic errors 6/129 (4.6%)**  **Total 6/129 (4.6%)** | - Necrosis/Metastasis  - Ependymoma/Pineocytoma  - Gliosis |  | - GBM  - Oligodendroglioma  - Astrocytoma III |
| Callovini, 2008 | USA  “In a series of 271 stereotactic brain biopsy performed from 1999 to 2005, 174 cases were selected in which clinical and radiological findings indicated high- or low-grade supratentorial glial lesions.” | N = 174  Age median 37  M:F 98:74 | **Diagnostic errors 11/107 + 10/67 = 21/174 (12.1%)**  **Misgraded 10/67 or in total 10/174 (5.7%)**  **Total:**  **31/174 (17.8%)** | HGG (107)  LGG (67) | - Anaplastic astrocytoma  (wrong grading) | - Lymphoma  - Demyelinating disease  - Metastatic tumour  - Infarction  - Neurocysticercosis  - Radionecrosis  - Inflammation  - Ischemia |
| Campos, 2009 | Germany  “Patients were collected from independent neurosurgery,epileptology, and neuropathology databases. Over a period of 20 years (1988–2008), 61 patients had been operated with subtotal or total DNT removal.” | N = 61  Age mean 27  M:F 27:34 | **Diagnostic errors 8/61 (13.11%)**  **Total: 8/61 (13.11%)** | - Astrocytoma WHO I  - Astrocytoma WHO II  - Oligodendroglioma WHO II |  | - Dysembryoplastic neuroepithelial tumor (DNT) |
| Ghosal, 2011* | India  “Total of 306 cases of intracranial lesions were evaluated preoperatively by smear preparation in the Department of Pathology from January 2003 to May 2007” | N = 306  Age range 3-63  M:F ratio 1.5:1 | **Diagnostic errors 7/306 (2.3%)**  **Misgraded**  **10/306 (3.3%)**  **Total 17/306 (5.6%)** | - Glioblastoma multiforme/ganglioglioma/pilocytic astrocytoma  - Ependymoma  - Ependymoma  - Oligodendroglioma  - Oligoastrocytoma/meningioma  - astrocytoma | - Anaplastic astrocytoma  - Pilocystic astrocytoma  - Glioblastoma multiforme  - Fibrillary astrocytoma  - Oligodendroglioma | - (Clear cell) ependymoma |
| Constantini, 2013* | 9 countries (India, Brazil, UK, Germany, Turkey, Japan, Italy, Israel, Canada)  “Retrospective data were collected from 13 randomly chosen medical centers that routinely performed NEBs (neuroendoscopic biopsy) between January 1998 and December 2009.” | N = 293  NEB and open surgery: N = 78  Age mean 28.3  M:F 176:117 | **Diagnostic errors 3/78 (3.8%)**  **Misgraded**  **7/78 (9.0%)**  **Total 10/78 (12.8%)** | - Low grade astrocytoma  - Low grade astrocytoma  - Low grade astrocytoma | - Ganglioglioma  - Juvenile pilocytic astrocytoma  - High grade astrocytoma | - Benign teratoma  - Cavernoma  - Meningioma |
| Yao, 2014 | China  “From September 2010 to August 2013, 47 consecutive image-guided stereotactic biopsies of parenchymal brain lesions were performed in. This was a retrospective study.” | N = 47  (trial group n = 24, control group n = 23)  Age median 49.7  M:F 31:16 | **Misgraded 4/24 (16.7%)**  **Total 4/24 (16.7%)** | - Astrocytoma II  - Anaplastic Astrocytoma III | upgraded  - Anaplastic Astrocytoma III  - Glioblastoma IV |  |
| Hamasaki, 2018* | USA  “A natural language search was undertaken in our CoPath database to analyse patients who underwent brain biopsy at the Queen’s Medical Center between January 1, 1996 and January 1, 2016” | N = 400  Neoplastic lesions n = 338  Age range 1-93  M:F ratio 1.1:1 | **Diagnostic errors 15/338 (4.4%)**  **Misgraded 2/338 (0.6%)**  **Total: 17/338 (5.0%)** | - Astrocytoma  - Astrocytoma/gliosis  - Lymphoma/nondiagnostic/necrotic tissue | - Oligoastrocytoma  - Oligodendroglioma  - Glioblastoma  - Ganglioglioma | - Metastatic lung carcinoma  - Subependymoma  - Hemangiopericytoma  - PNET  - DNT |
| Maldonado, 2018 | USA  “Cases and controls were accrued from an institutional neuro-oncology/neuroradiology database containing a total of 304 diffuse gliomas.” | N = 304  Age median 54  M:F 6:12  (age and sex from the 18 misgraded patients) | **Misgraded**  **18/304 (5.9%)**  **Total: 18/304 (5.9%)** | Diffuse Glioma | - Glioblastoma  - Diffuse Astrocytoma  - Oligodendroglioma (anaplastic)  - Oligoastrocytoma  - |  |
| McCullough, 2018 | USA  “We identified patients presented during our routine tumor board conference with a new diagnosis of a diffuse infiltrating glioma, WHO grades II-IV, between January 1, 2007 and December 31, 2014” | N = 146  Patients who underwent resection: N = 13  Age mean 53  M:F 80:66 | **Misgraded 3/13 (23%)**  **Total: 3/13 (23%)** | Glioma WHO Grade II - IV | Upgraded:  AA - GB  LGA - AA |  |
| Obeidat, 2019* | Jordan  “This retrospective study comprised all brain tumor cases with FS and permanent section diagnoses from July 1, 2007 to December 31, 2017 at Jordan University Hospital in Amman, Jordan.” | N = 179  Age mean 44.7  M:F 84:95 | **Diagnostic errors 5/179 (2.8%)**  **Misgraded 7/179 (3.9%)**  **Total: 12/179 (6.7%)** | - Small round blue cell tumor/Metastasis/Medulloblastoma  - Gliosis  - Glioblastoma | - Low grade Glioma  - Anaplastic Astrocytoma  - Glioblastoma  - High Grade Glioma  - Astrocytoma II | - Glioblastoma  - Anaplastic Oligodendroglioma -(Astroblastoma)  -(Pleomorphic xanthoastrocytoma)  - (Pilocytic Astrocytoma)  - Metastatic carcinoma |
| Zoccarato, 2019 | Italy  “Patients with LE were selected retrospectively from the cohorts of three Italian neurological centers specialised in the recognition and treatment of autoimmune neurological diseases.” “Patients affected by temporal tumors were selected retrospectively from the database of the Istituto Oncologico Veneto (Padua). A total of 749 patients were screened from April 1998 to August 2017.” | N = 54  (Limbic encephalitis n = 25, tumour group n = 24  M:F 29:25  Age mean 59 | **Diagnostic errors 2/22 (9.1%)**  **Total: 2/22 (9.1%)** | - Limbic encephalitis |  | - Glioma |
| Sasagawa, 2021 | Japan  “We retrospectively reviewed pathological records for lesions that were a brain tumor. In total, 557 patients received brain tumor removal at Sapporo Medical University between January 2011 and September 2019.” | N = 214  Age median 55  M:F 115:99 | **Diagnostic errors 12/224 (5.4%)**  **Total: 12/224 (5.4%)** | - Ischemic stroke (= stroke mimic)  - Glioma |  | - Glioblastoma/glioma  - Hemorrhagic infarction/stroke |
| Yu, 2023* | China  “Between July 2017 and September 2021, 397 consecutive patients whose histopathological diagnoses were confirmed as GBM (*n* = 266) or PCNSL (*n* = 131) were identified. Among these, 239 patients were excluded due to multiple reasons” | N = 158  (GBM n = 70, PCNSL n = 88)  Age median GBM = 52  PCNSL = 54.5  M:F GBM= 43:24  PCSNL 54:34 | **Diagnostic errors 5/88 (5.7%)**  **Total: 5/88**  **(5.7%)** | Glioblastoma multiforme |  | -PCNSL (primary central nervous system lymphoma) |

In order of publication year. For the columns "Country and patient selection" and " we use the exact wording provided by the original study authors. Note that we report the diagnoses as provided by the study authors in the original manuscript (even if some diagnostic concepts might be outdated or refer to broad diagnostic classes). Studies where the initial glioma diagnosis was based on histology are marked with an *.

*Abbreviations: CNS, central nervous system; CT, computed tomography; DACI, Desmoplastic astrocytoma of infancy; DIGG, desmoplastic infantile ganglioglioma; DNT, Dysembryoplastic neuroepithelial tumor; FS, frozen section; GBM, glioblastoma multiforme; HGG, high grade glioma; LE, limbic encephalitis; LGG, low grade glioma; MRI, magnetic resonance imaging; NEB, neuroendoscopic biopsy; PXA, pleomorphic xanthoastrocytoma; PCSNL, primary central nervous system lymphoma; PNET, Primitive neuroectodermal tumor; SB, stereotactic biopsy.*
